# Genetic correlation and causal relationships between cardio-metabolic traits and Lung function Impairment

**DOI:** 10.1101/2020.09.16.20190306

**Authors:** Matthias Wielscher, Andre F.S. Amaral, Diana van der Plaat, Louise V. Wain, Sylvain Sebert, David Mosen-Ansorena, Juha Auvinen, Karl-Heinz Herzig, Abbas Dehghan, Debbie L Jarvis, Marjo-Riitta Jarvelin

## Abstract

**Background:** Associations of low lung function with features of poor cardio-metabolic health have been reported. It is, however, unclear whether these co-morbidities reflect causal associations, shared genetic heritability or are confounded by environmental factors.

**Methods:** We performed three analyses: 1) cardio-metabolic health to lung function association tests in NFBC1966, 2) cross trait LD score regression to compare genetic backgrounds and 3) Mendelian Randomization (MR) analysis to assess the causal effect of cardio-metabolic traits and disease on lung function, and vice versa (bidirectional MR). Genetic associations were obtained from UK Biobank data or published large-scale genome-wide association studies (N > 82,000).

**Results:** We observed negative genetic correlation between lung function and cardio-metabolic traits and diseases. In Mendelian Randomisation analysis (MR) we found associations between Type 2 Diabetes instruments and FVC as well as FEV1/FVC. BMI instruments were associated to all lung function traits and CRP instruments to FVC. These genetic association provide evidence for a causal effect of cardio-metabolic traits on lung function. Multivariable MR suggested independence of these causal effects from other tested cardio-metabolic traits and diseases. Analysis of lung function specific SNPs revealed a potential causal effect of FEV1/FVC on blood pressure.

**Conclusions:** The present study overcomes many limitations of observational studies by using Mendelian Randomisation. We provide evidence for an independent causal effect of T2D, CRP and BMI on lung function with some of the T2D effect on lung function being mediated by CRP. Furthermore, this analysis suggests a potential causal effect of FEV1/FVC on blood pressure. Our detailed analysis of the interplay between cardio-metabolic traits and impaired lung function provides the opportunity to improve the quality of existing intervention strategies.

## Background

Obesity and metabolic syndrome have become an increasing public health problem in most parts of the world. By 2025, global obesity prevalence is predicted to reach 18% in men and 21% in women (1). The associations of obesity with chronic non-communicable diseases such as type 2 diabetes, cardiovascular disease and cancers are well described. Meanwhile there is a growing literature on the association of obesity with lung function and chronic lung disease, although the underlying pathways and potential mediators are not well understood.

Several observational studies have reported an association between low lung function and metabolic syndrome traits, including obesity (2-5). In this report we replicated these associations using data from a population-based cohort, the Northern Finland Birth Cohort (NFBC1966). However, the associations cannot be translated to causation since observational studies are not able to control for all known potential confounders, residual confounding and reverse causation. However, it is not possible to infer whether associations such as those seen in NFBC1966, and in the other studies, are causal as most studies were not able to control for all known potential confounders or residual confounding by unknown factors.

In this study we assess associations between eleven cardio-metabolic traits representing wider range of traits than usually accounted in pure metabolic syndrome definition (6, 7) Body mass index (BMI)(8), Type 2 Diabetes (T2D)(9), C-reactive protein (CRP)(10), high density lipoprotein (HDL), low density lipoprotein (LDL), total cholesterol (TC), triglycerides (TG)(11), diastolic blood pressure (DBP), systolic blood pressure (SBP), pulse pressure (PP)(12), coronary artery disease (CAD)(13), and three lung function outcomes (First second forced expiratory volume (FEV1), Forced Vital Capacity (FVC) and a ratio of both FEV1/FVC). We examine whether these metabolic traits and lung function are genetically correlated using a cross trait linkage disequilibrium (LD) score regression and then go on to determine whether the associations are likely to be causal, using Mendelian Randomisation (MR). MR is a method to estimate causal effects by using genetic variants with known effects on the risk factor of interest as a proxy (i.e. instrumental variable) (14) and, as long as underlying assumptions are not violated (15, 16), MR is not susceptible to classical confounding (as seen in observational studies) or reverse causation (Figure 1A).

**Figure 1:**
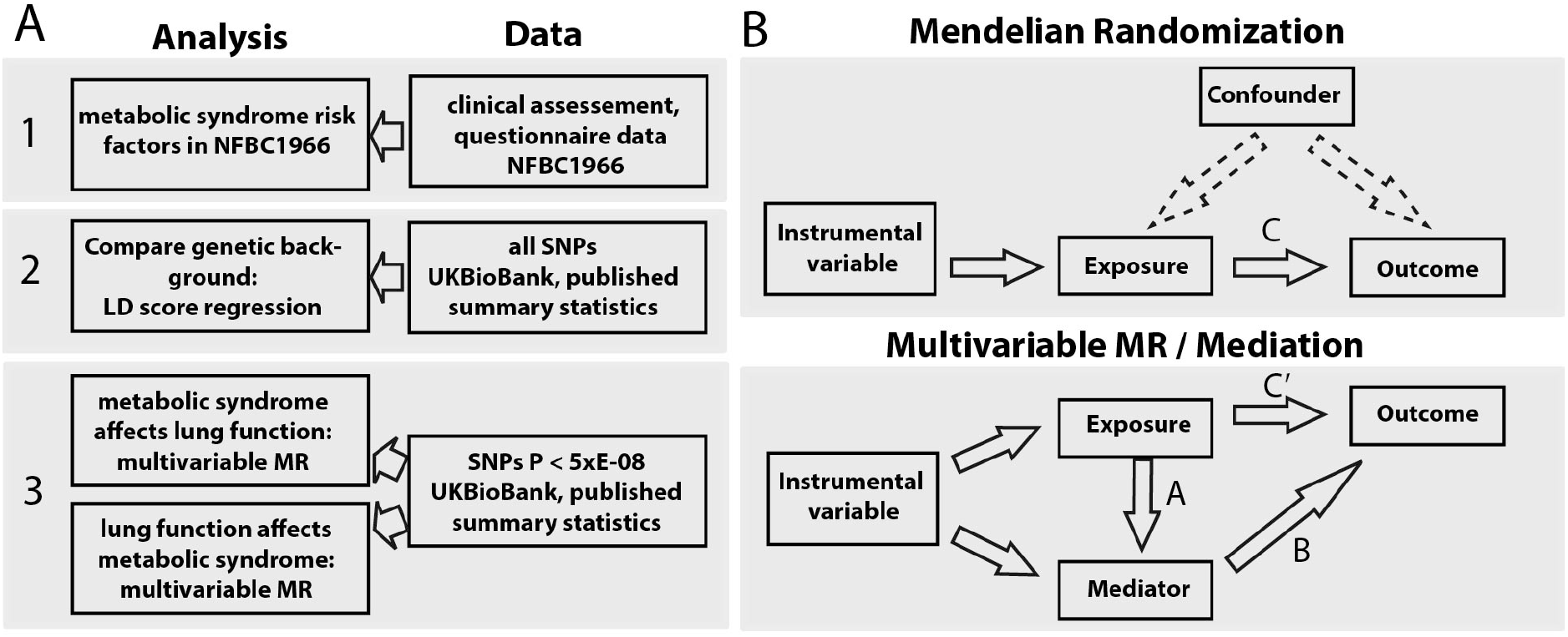
Flow chart of study design. A, Cardio-metabolic traits were Body mass index (BMI), Type 2 Diabetes (T2D), C-reactive protein (CRP), Lipoprotein (HDL-C), low density lipoprotein (LDL-C), Total cholesterol (TC), Triglycerides (TG), Systolic Blood pressure (SBP), Diastolic blood pressure (DBP), pulse pressure (PP) and coronary artery disease (CAD). Tested lung function traits were First second forced expiratory capacity (FEV1), Forced Vital Capacity (FVC) and a ratio of both FEV1/FVC. B. Graphical diagrams relationships in a classical MR and Mediation analysis. Upper panel gives overview of MR analysis, indicating the use of genetic instruments instead of the actual exposure. C in upper panel refers to the causal estimate as well as the C-path in Mediation analysis setting. Lower panel gives overview of mediation analysis following Baron Kenney approach. For mediation analysis in this study we subtracted C’ path from C path to get effect sizes for mediation.

Mendelian Randomisation has been used to estimate the causal effect of body mass index (BMI) on two lung function parameters (forced expiratory volume in one second, FEV1, and forced vital capacity, FVC) (17). However, it did not account for potential shared genetic instruments (in this case whether the genetic instruments modify lung function through factors other than BMI). To overcome this problem, we used a wide range of MR methods, amongst others a recently developed extension of MR, multivariable MR (MMR) (18), which models the effects of pleiotropy, estimating the independent causal effects of each risk factor simultaneously. This analysis setting allowed us to take advantage of horizontal pleiotropy and to gain insights into the interplay between cardio-metabolic traits through the comparison of MMR estimates and univariable MR estimates of each risk factor (Mediation Analysis). Moreover, former papers did not test for potential causal effects in the opposite direction as we do here by conducting a bidirectional MR (Figure 1B).

## Methods

### Studied traits

In this study we assessed associations between eleven cardio-metabolic traits and three lung function outcomes (FEV1, FVC and the ratio of FEV1/FVC). Cardio-metabolic traits are: Body mass index (BMI)(8), Type 2 Diabetes (T2D)(9), C-reactive protein (CRP)(10), four blood lipid levels outcomes (high density lipoprotein (HDL-C), low density lipoprotein (LDL-C), total cholesterol (TC), and triglycerides (TG)(11), three blood pressure outcomes (diastolic blood pressure (DBP), systolic blood pressure (SBP), and pulse pressure (PP))(12), coronary artery disease (CAD)(13), and three lung function outcomes (FEV1, FVC and the ratio of FEV1/FVC).

### Observational associations with lung function in NFBC1966

The Northern Finland Birth Cohort 1966 (NFBC1966) has been described in detail elsewhere. Associations of the cardio-metabolic traits and lung function outcomes were determined in 4968 participants with lung function and information on cardio-metabolic traits of interest as well as relevant confounders at participants age of 46 years (see Additional File 1 methods for more details).

### Associations of SNPs with cardio-metabolic traits

We extracted the effect estimates for SNPs associated (P< 5×10^-8^) with the cardio-metabolic traits from the most recent published GWAS including 82,000 up to 322,000 individuals (Table 1, Table A in Additional File 1, Table 2 in Additional File 2)(8-11, 13, 19). We used the SNP with the lowest P-value within a genomic region of 1 mega base as the instrument.

**Table 1:**
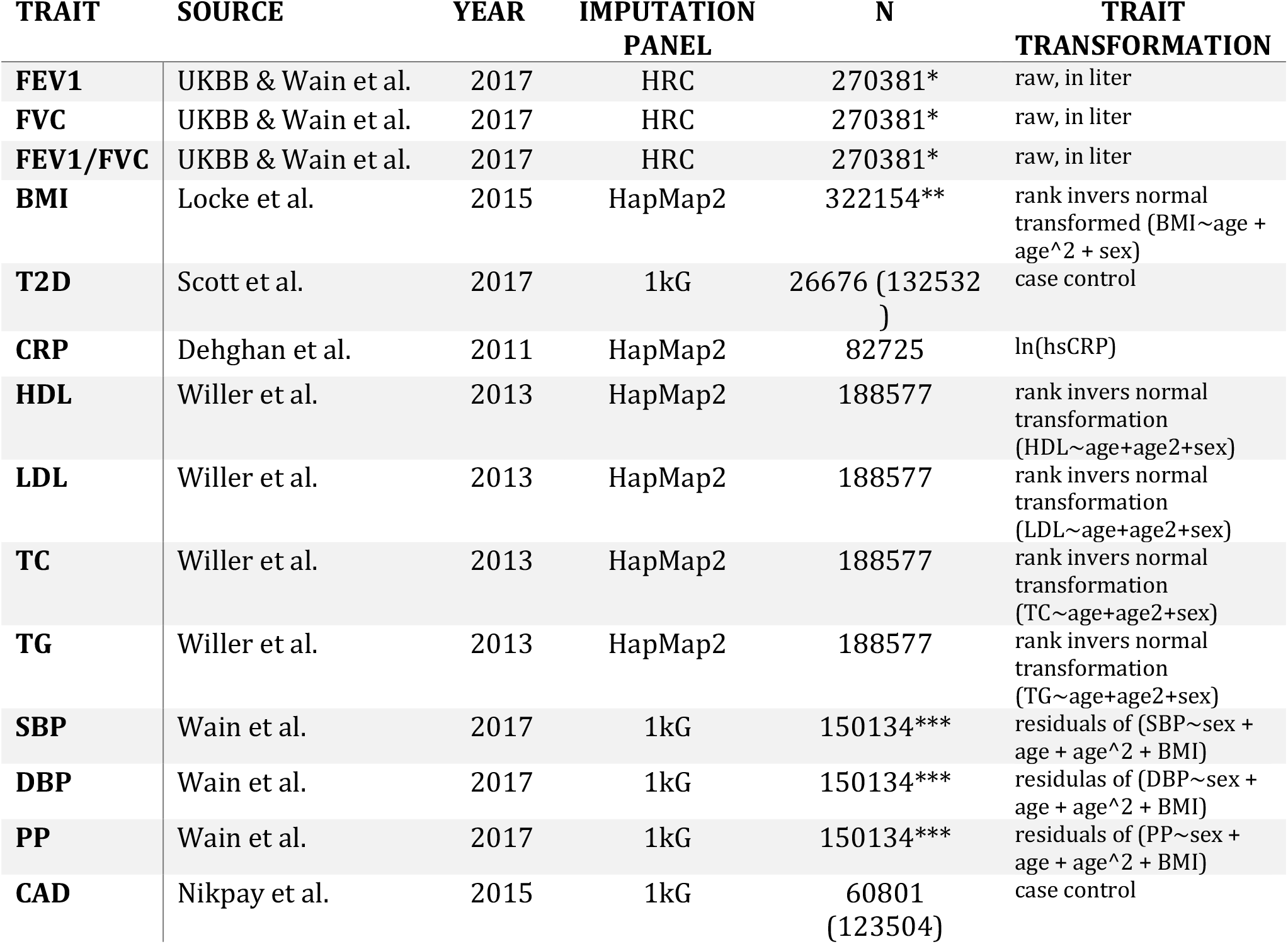
Data used for the Mendelian randomization analysis. For CAD and T2D participant numbers were split into cases and controls. *Reproducibility of spirometry measurement using ERS/ATS criteria; ** European ancestry; *** stage1 meta-analysis

| TRAIT | SOURCE | YEAR | IMPUTATION<br>PANEL | N | TRAIT<br>TRANSFORMATION |
| --- | --- | --- | --- | --- | --- |
| **FEV1** | UKBB & Wain et al. | 2017 | HRC | 270381* | raw, in liter |
| **FVC** | UKBB & Wain et al. | 2017 | HRC | 270381* | raw, in liter |
| **FEV1/FVC** | UKBB & Wain et al. | 2017 | HRC | 270381* | raw, in liter |
| **BMI** | Locke et al. | 2015 | HapMap2 | 322154** | rank invers normal transformed (BMI~age + age^2 + sex) |
| **T2D** | Scott et al. | 2017 | 1kG | 26676 (132532) | case control |
| **CRP** | Dehghan et al. | 2011 | HapMap2 | 82725 | ln(hsCRP) |
| **HDL** | Willer et al. | 2013 | HapMap2 | 188577 | rank invers normal transformation (HDL~age+age2+sex) |
| **LDL** | Willer et al. | 2013 | HapMap2 | 188577 | rank invers normal transformation (LDL~age+age2+sex) |
| **TC** | Willer et al. | 2013 | HapMap2 | 188577 | rank invers normal transformation (TC~age+age2+sex) |
| **TG** | Willer et al. | 2013 | HapMap2 | 188577 | rank invers normal transformation (TG~age+age2+sex) |
| **SBP** | Wain et al. | 2017 | 1kG | 150134*** | residuals of (SBP~sex + age + age^2 + BMI) |
| **DBP** | Wain et al. | 2017 | 1kG | 150134*** | residuals of (DBP~sex + age + age^2 + BMI) |
| **PP** | Wain et al. | 2017 | 1kG | 150134*** | residuals of (PP~sex + age + age^2 + BMI) |
| **CAD** | Nikpay et al. | 2015 | 1kG | 60801 (123504) | case control |

### Associations of SNPs with lung function

We obtained effect estimates of the selected cardio-metabolic SNPs on lung function (FEV1, FVC and FEV1/FVC) from UK Biobank (UKB; Application Number 19136) using BOLT-LMM adjusted for assessment centre, sex, age, height, current smoking status and pack-years (See Table B in Additional File 1 for UKB characteristics).

### Cross trait LD score regression

We assessed the genetic correlation between each metabolic trait and each lung function parameter using the recommended settings in the software package LDSC (v1.0.0)(20). Briefly, this method generates a score reflecting whether the GWAS test statistic of a biologically relevant variant correlates with nearby variants in high linkage disequilibrium. The z statistic for the genetic association of each variant with trait 1 are multiplied with the z statistic for the genetic association with trait 2, followed by regression of this product of statistics against the LD scores. The slope (coefficient) represents genetic correlation. When large, the same genetic variants impact both the traits.

### Mendelian Randomization (MR)

We performed a 2-sample Mendelian Randomization, using the CRAN package *Mendelianrandomization*, unless stated otherwise (Figure 1B). We excluded instruments having a direct effect on the outcome (P<5xE-08) and palindromic SNPs. We estimated the causal effect of a single risk factor on lung function using the widely used fixed-effect IVW MR. Further, we performed sensitivity analyses using weighted median, mode based and MR Egger methods to rule out potential pleiotropy. To further assess the stability of our results we used penalized MR approaches and reproduced the results using altered sets on input SNPs. We achieved this via exclusion of critical variants as suggested by MR-PRESSO(21) and contamination mixture method(22) (Additional File 1 methods, Table 3 in Additional File 2)

#### Multivariable MR (MMR)

To assess the independent effects of each cardio-metabolic trait, while accounting for the effects of the others, we used multivariable MR (18) (Additional File 1 methods, Table 4 in Additional File 2),. We regressed the coefficients for the SNP-outcome association against all risk factors separately and then simultaneously for each risk factor. The residuals of these regressions were used as the outcome to estimate the causal effect.

#### Mediation analysis

We tested traits as mediators when they were consistently significantly (p<0.05) associated with lung function in the univariable MR. For those traits, we compared the direct effect estimates (IVW MR of risk factor) with the total effect estimate (MMR estimate of risk factor plus Mediator)(23)(Figure 1B,).

#### Bidirectional MMR

We repeated the MR analysis in the opposite direction to determine possible causal effects of lung function on cardio-metabolic traits (Figure 1B). For this we used a set of validated SNPs described by Wain et al (19) as instruments for lung function. (Additional File 1 methods, Table A in Additional File 1).

## Results

Cardio-metabolic traits are closely related and correlation across traits could generate horizontal pleiotropy. For example, if an instrument for trait A is also associated with trait B (e.g. variants in FTO for BMI and CRP), it would be challenging to find out whether the association with outcome (e.g. lung function) is reflecting the causal effects of trait A or B. This is a major challenge in this study as 17% of the variants we used as instruments in this MR are associated with more than one cardio-metabolic trait (Table 2 in Additional File 2). To address this, we use outlier robust methods such as weighted median MR and Mode based estimation (Figure 3) also we performed sensitivity analysis on altered sets variants for each exposure (Additional File 1 methods, Table 2 in Additional File 2). Finally, by adding every risk factor to our MR model separately (MMR), we evaluate the independence of the tested effect as well as the attenuation as mediated fraction of the added risk factor on the outcome (Figure 4).

In the following we present results of the three analyses for each cardio-metabolic trait (Figure 1):

1. Observational analyses of the association between traits, using the data from the NFBC1966 study (Table C-D in Additional File 1).
2. Evidence for genetic correlation using cross-trait LD score regression (Figure 2, Table E in Additional File 1).
3. Evidence for causal associations from Mendelian Randomization analysis of cardio-metabolic traits on lung function, and vice-versa (Figure 3, Figures A-F in Additional File 1, Table S3). Robust associations between instruments of the risk factors and outcomes were followed up by Multivariable MR (Figure 4). This allowed us to draw conclusions if any potential causal effects were independent from other tested risk factors (horizontal pleiotropy) and if we can find certain proportions of the effect being mediated by other cardio-metabolic traits.

**Figure 2:**
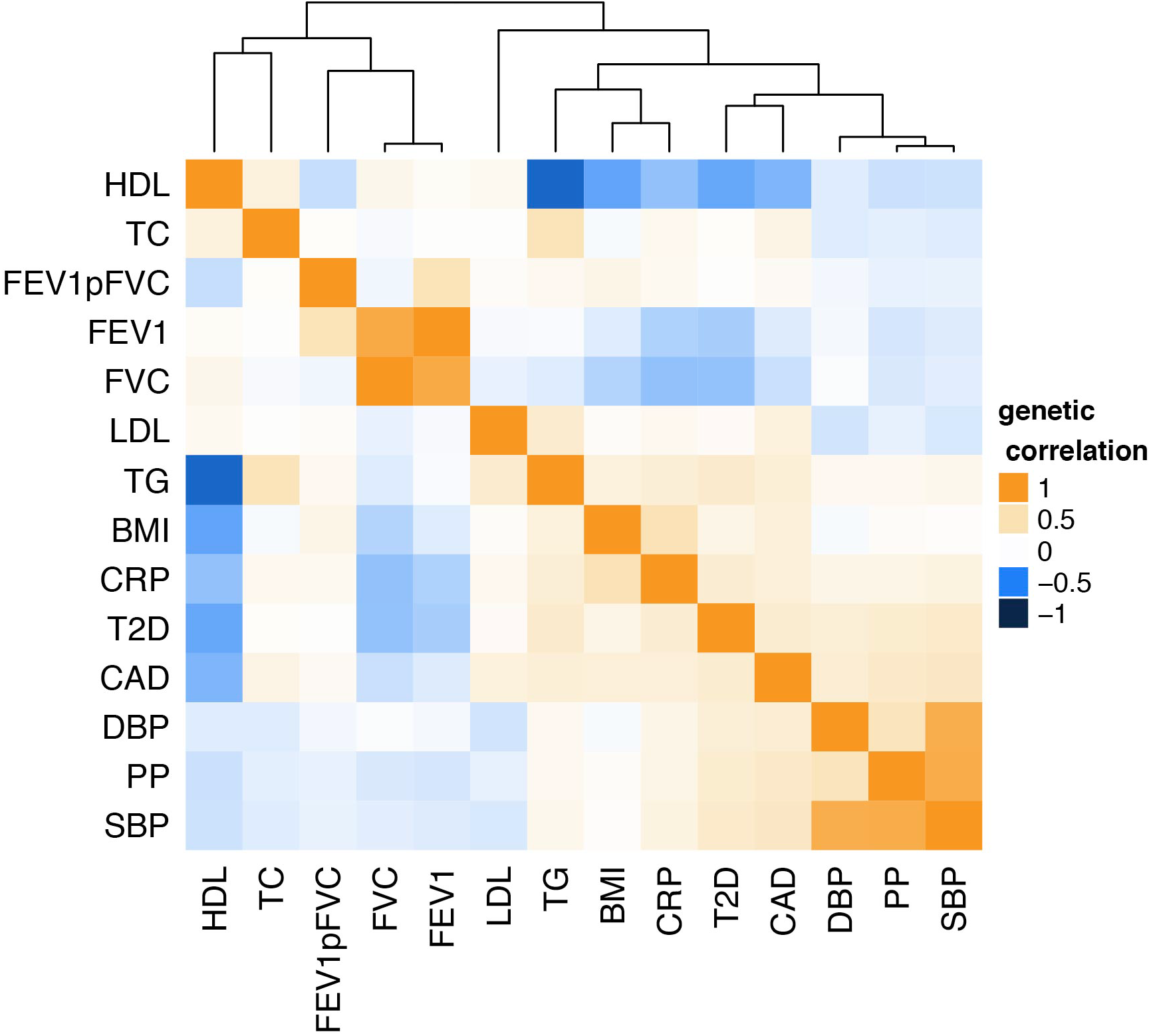
Heat map of genetic correlation (Cross trait LD score regression). Blue boxes indicate negative correlation; orange boxes positive genetic correlation. Distance on cluster dendrogramm measures the similarity between traits. Correlation values including P-values between lung function traits and cardio-metabolic traits are given in table F in Additional File 1.

## BMI

We found negative associations of BMI with FEV1 and FVC, and positive associations with FEV1/FVC ratio in the observational analyses in NFBC1966. Consistent with this, cross trait LD score regression showed the same direction of effects (FEV1/FVC P=1.8×E-13, FEV1 P=8×E-04, FVC P=9.9×E-18; Figure 2). For Mendelian Randomization analysis we used BMI specific SNPs described in Locke et al.(8) F-statistic of variants was 27 or higher (Table 2 in Additional File 2). We note that 12 SNPs deployed as BMI instruments also reach genome wide significance for one or more of the tested cardio-metabolic traits (Table 2 in Additional File 2). Evaluation of several MR approaches suggests a causal effect of BMI on all lung function parameters (Figure 3A). Effect sizes derived from the IVW MR analysis in UK Biobank data showed a decrease of 24ml in FVC and 12ml of FEV1 per unit (kg/m2) change in BMI. Low P values obtained from multivariable MR (MMR) suggest that the effects of BMI on lung function are independent from other tested risk factors (Figure 4). However, we observed strong attenuation of BMI effects on FVC and FEV1/FVC when adding genetic instruments for diastolic blood pressure, triglycerides or HDL-C to multivariable MR model (Figure 4, Table 5 in Additional File 2). Multivariable MR also suggests that 2 % of the BMI effect on restrictive lung patterns (indicated by lower FVC values) is mediated by CRP.

### Type 2 Diabetes

As seen for BMI, T2D was negatively associated with FEV1 and FVC and positively associated with FEV1/FVC lung patterns in NFBC1966 (Table D in Additional File 1). Cross trait LD score regression showed a negative genetic correlation with both FVC (P=9.2×E-13) and FEV1 (P=3.1×E-10) and a positive but statistically non-significant Bonferroni corrected genetic correlation with FEV1/FVC (P=0.015 Figure 2). We used genetic instruments for T2D described in Scott et al.(9) (table 1). All variants had an F statistic above 30 (Table 2 in Additional File 2). Eleven T2D instruments were associated to one or more of the other tested risk factors in this study. Applying several MR techniques, we found a consistent association between T2D specific SNPs and FVC and FEV1/FVC (Figure 3). Effect sizes derived from the IVW MR analysis suggested a decrease of 65 ml in FVC and 108 ml of FEV1 with the presence of T2D. P values obtained from Multivariable MR indicate the effect of T2D on impaired lung function is independent from most tested risk factors. We observed strong attenuation of the T2D effect on FVC when adding instruments for SBP or PP (Figure 4) to multivariable regression model. Adding genetic instruments for CRP to the model shows that CRP mediates 14.9% of the T2D effect on FVC. There was no evidence that BMI mediated the effect on T2D on FVC or FEV1/FVC (Figure 4).

**Figure 3:**
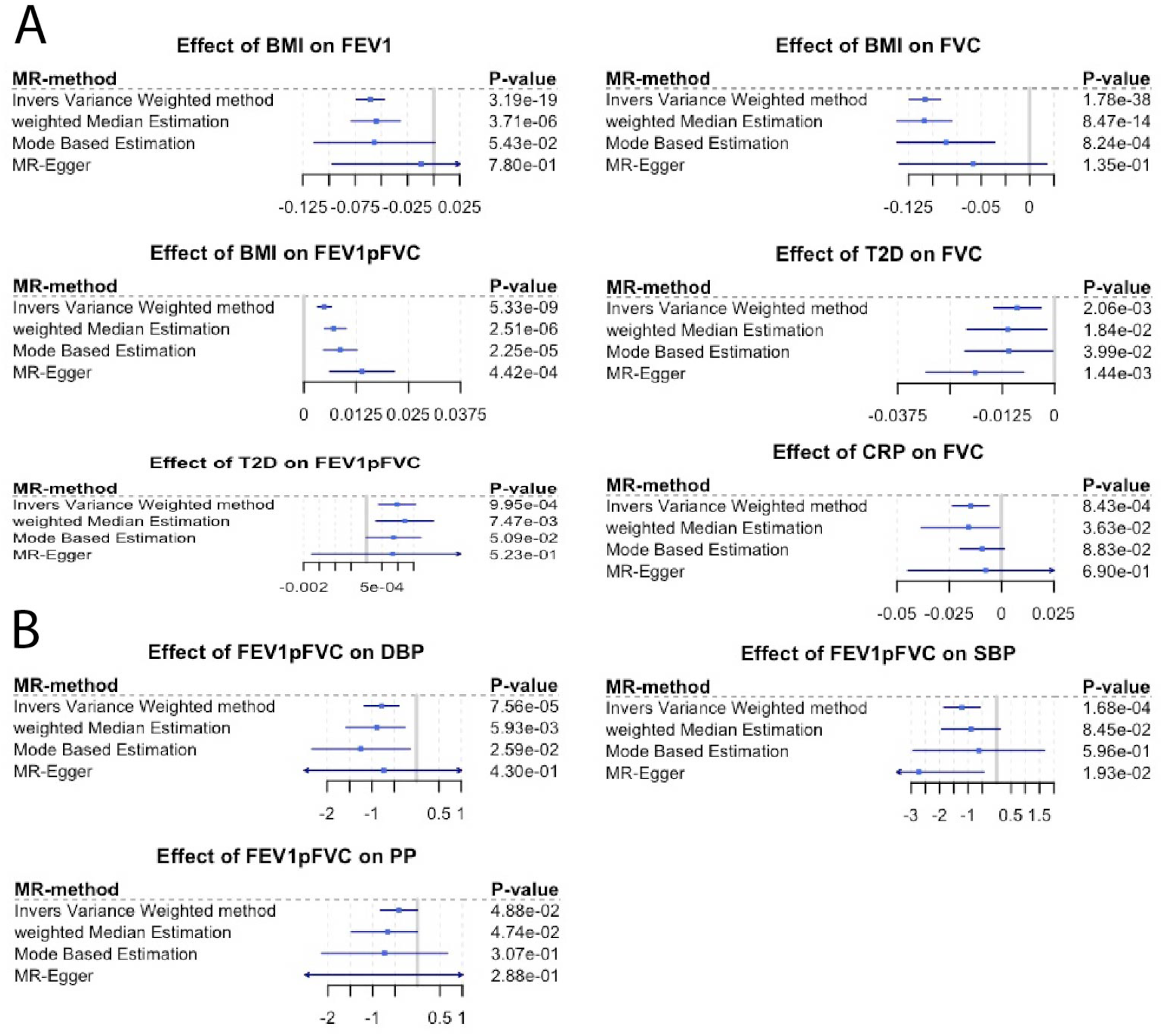
Forest plot of Mendelian Randomization result. Blue square represents causal estimate. Blue line is 95% confidence interval. Every line represents one approach to estimate the potential causal effect (Additional File 1 methods). Section A represents effects of tested risk factors on impaired lung function. Section B is the invers direction. Impaired lung function as exposure for blood pressure. If causal effect estimates were not nominal significant with at least two different approaches and did not have a consistent direction of effect they are given in Figures A-F in Additional File 1

## CRP

Within NFBC, we found a strong negative association between blood CRP levels and all three lung function parameters (Table D in Additional File 1). There was strong evidence of a genetic correlation between lung function and blood CRP levels (FEV1/FVC P=0.0528, FEV1 P=4.29E-06, FVC P=1.15E-11; Table E in Additional File 1, Figure 2). We used CRP instruments described in Dehghan et al (table 1)(10). We note that seven out of the 18 CRP instruments were associated with one or more cardio-metabolic trait (Table 2 in Additional File 2). Genetic instruments for CRP were significantly associated with FVC, suggesting a causal effect of CRP on restrictive lung patterns (Figure 3). We observed a decrease of 14ml in FVC per log change in serum CRP level. MMR analysis supports a causal effect of CRP on FVC (Figure 4) with strong attenuation of the effect when adding total cholesterol, LDL-C or HDL-C to the model. Furthermore, multivariable MR analysis shows that 8.3% of the CRP effect on FVC is mediated by BMI.

### Lipid levels

HDL cholesterol and triglyceride levels were associated with lung function in NFBC1966 (Figure 3). There was a negative genetic correlation of triglycerides with FVC (P=0.029) but a positive correlation with FEV1/FVC (P=0.012), with the opposite for HDL cholesterol (positive correlation with FVC, P=3.8×E-03; negative correlation with FEV1/FVC, P=4.5×E-04, Table E in Additional File 1). For LDL-C and total cholesterol no significant associations were seen in the observational data nor was there evidence of genetic correlation. We did not see consistent associations between any lipid trait and lung function in the MR analyses (Figures A-C in Additional File 1).

### Blood pressure

We observed a negative association of both DBP and SBP with FEV1 and FVC and a positive association with FEV1/FVC (Table D in Additional File 1, Figure 3) in NFBC1966, but no associations with PP. LD score regression showed negative genetic correlation between FEV1 and PP (P=1.8×E-03, Figure 2, Table E in Additional File 1) only. We did not observe consistent associations between any blood pressure trait and lung function in MR analyses (Figures A-C in Additional File 1)

### Coronary artery disease

A diagnosis of CAD was not associated with lung function in NFBC1966. (Appendix table 4). Cross trait LD score regression showed negative correlation with FEV1 and FVC, but a positive correlation with FEV1/FVC (FEV1/FVC P=8.2xE-03, FEV1 P=2xE-03, FVC P=3.1xE-06, Figure 2). MR suggested these correlations were not causal. (Figures A-C in Additional File 1).

**Figure 4:**
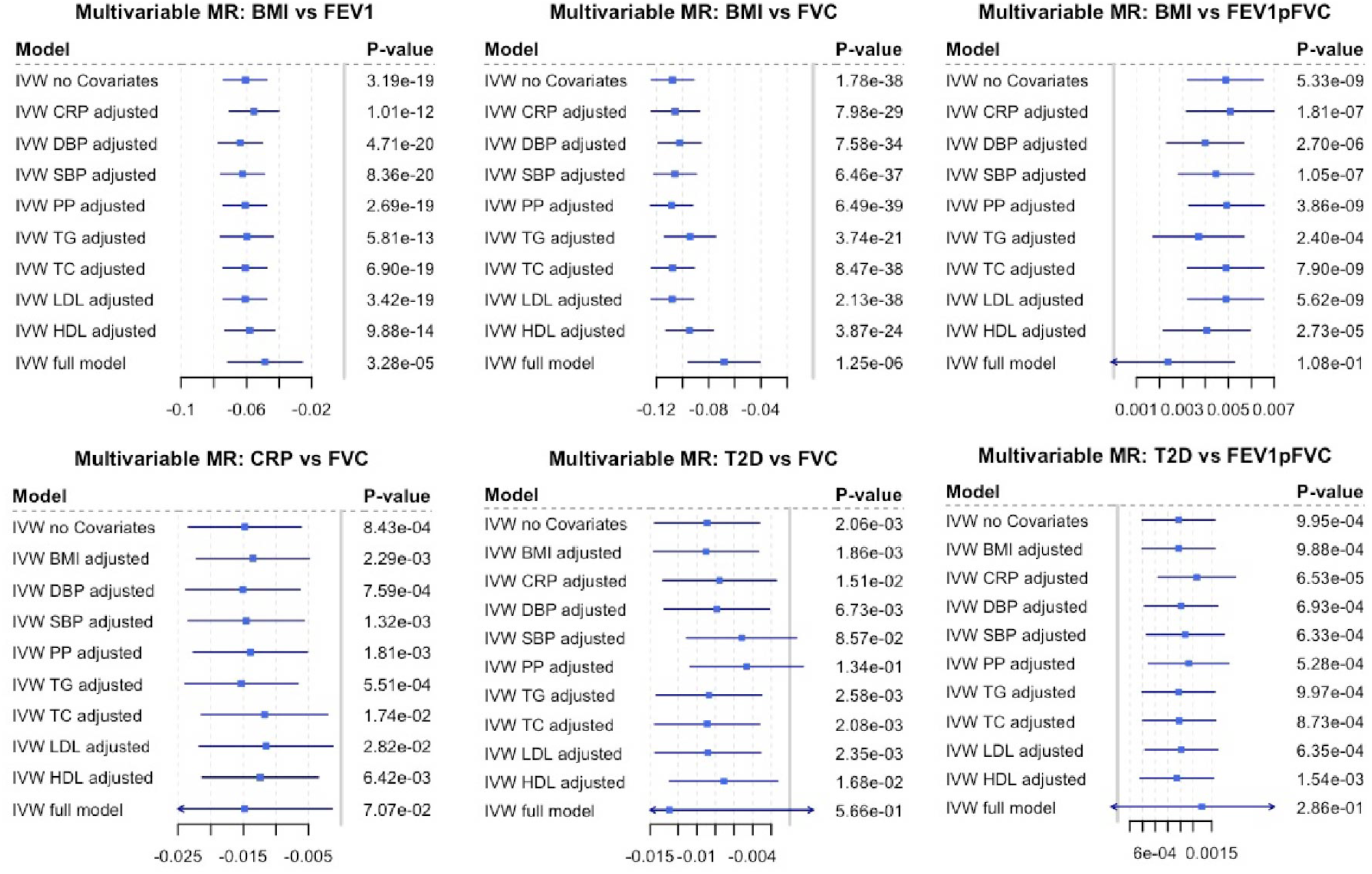
Multivariable MR and Mediation analysis. First entry in each plot is the inverse variance weighted causal estimate as given in Figure 3. This estimate represents the direct effect of the risk factor on the outcome. Subsequent lines are adjusted for one risk factor each representing the total effect. Full model has all risk factors as covariate in the model. Differences in effect sizes resulting from attenuation can be interpreted as mediated by the exposure added to the model.

### Causal effect of lung function on cardio-metabolic traits

We used variants described by Wain et al.(19) as instruments (table 1) to test for possible causal effects of lung function on cardio-metabolic traits. We discovered consistent associations between FEV1/FVC specific SNPs and DBP, SBP as well as PP suggesting a causal effect of FEV1/FVC on blood pressure (Figure 3B, Figures D-F in Additional File 1).

## Discussion

In summary, our findings suggest that lung function parameters are genetically correlated with multiple cardio-metabolic traits (BMI, T2D, CRP, HDL-C, LDL-C, TC, TG, SBP, DBP, PP, CAD). Furthermore, we found evidence for causal effect of some cardio-metabolic traits (BMI, T2D, CRP) on lung function measures and a possible causal effect of FEV1/FVC on blood pressure. As the assessed cardio-metabolic traits were highly correlated, we used Multivariable Mendelian Randomization (MMR) to validate the findings from univariable MR and investigate the interplay between cardio-metabolic traits as the method simultaneously accounts for multiple causal factors (Figure 1B, Figure 4).

We have used state of the art statistical methods for assessing genetic correlation, using publicly available summary statistics of genetic associations. This method has been previously used to assess genetic correlations for example between 24 traits (including metabolic syndrome traits, mental health disorders, inflammatory bowel disease and educational attainment but not respiratory conditions) (20), between thirteen growth and eleven immune phenotypes (including asthma) (24), and between six cancers (including lung) and 14 non-cancer diseases (not respiratory conditions) (25). To our knowledge this method has not been used to report genetic correlations between the metabolic syndrome traits and lung function measures as shown here. One study has reported nominally significant genetic correlation with COPD for resting heart rate and hypertension (considered as a binary measure), with no correlation seen between COPD and stroke and other blood pressure traits (26).

We went on to determine whether the observed correlations could reflect causal associations using MR. LD score regression is not the same as Mendelian randomization as it uses information from the whole genome, and thus does not model one trait as a function of the other. Also, it makes no assumption of the causal direction of association (whereas MR tests the effect of one factor on the other). We show through MR that higher BMI is causally associated with lower FEV1 and FVC, with greater effects on the latter, explaining its positive effect on the derived parameter FEV1/FVC. This is consistent with previous reports from multiple large-scale population based epidemiological studies (27), as well as, with observational results from our analyses in the NFBC1966. Our Mendelian randomisation confirms these associations are very unlikely to be related to confounding by lifestyle factors related to BMI and lung function measures. Some have hypothesised that these associations could reflect reverse causation e.g. people with reduced lung function may be less likely to engage into physically activity and may subsequently gain weight. However, our bidirectional MR did not support this hypothesis.

Our analysis showed stronger effects of increased BMI on restrictive ventilation patterns than on airway obstruction. The mechanisms for how BMI affects lung function remain elusive although it is likely that fat accumulation between the muscles around the lungs and in the abdomen may have mechanical effects on the diaphragm and impede full inspiration as well as decreasing chest wall compliance (2). Obesity is also associated with increased levels of circulating pro-inflammatory markers such as CRP, IL6, TNFalpha and other cytokines(28). Systemic inflammation may explain some of the associations of obesity with impaired lung function. We indeed found that a small proportion (2%) of the effect of BMI on FVC was mediated by CRP, and 8.8% of the BMI effect on FEV1 was mediated by CRP (Figure 1B, Figure 4) suggesting indirect effects of BMI on airway obstruction through systemic inflammation.

Cross-sectional and longitudinal studies have shown that middle aged adults with T2D have worse lung function and slightly increased lung function decline (29). The proposed mechanisms are glycosylation of collagen within the lung, decreased muscle strength, impacts on surfactant proteins and low grade inflammation (5). Our MR supports the epidemiological observations confirming that T2D causally affects lung function (Figure 3A), independent of associations with BMI. We also observed low grade inflammation playing a key role in T2D lung function relationship as 14.9% of the T2D effect on FVC is mediated by CRP (Figure 4).

Similar to BMI we observed the causal effect of T2D having restrictive effects on the lung rather than obstructive (Figure 3). These associations persist when accounting for other cardio-metabolic traits. However, we do see an attenuation of the T2D effect on FVC when adding systolic blood pressure or pulse pressure to the MMR model.

Some longitudinal observational studies have reported that low lung function is an independent predictor of incident T2D (29, 30) and there is evidence that even in non-diabetics higher fasting glucose is associated with lower lung function (31). We found no evidence of causal associations of lung function on T2D, but the presence of low lung function in ‘prediabetics’ has been postulated to be related to lifestyle or environmental factors in utero, early childhood or adolescence that predispose individual to increased risks of diabetes and low lung function in the future (31). Our analysis is unable to address this hypothesis further.

We observed statistically significant effects of CRP on FVC (Figure 3). These associations were attenuated, but remained significant, after adjustment for BMI, pulse pressure or total cholesterol (Figure 4). This implies that CRP may contribute to impaired lung function as shown by many epidemiological studies(3, 4, 32).

Observational studies of associations of lipid levels and the presence of COPD have been inconsistent and a recent meta-analysis found no evidence for associations of COPD with serum levels of HDL-C, LDL-C, TC and TG (33). In cross-sectional studies any association may be masked by lipid-lowering treatments – and there may be underlying associations of COPD with high triglyceride levels (33). A large population based cross-sectional study in France showed strong associations of restrictive lung function deficits with cardio-metabolic, reporting associations with lipid profile (2). Our analysis does not support a causal effect of total cholesterol and triglycerides on lung function (Figures A-C in Additional File 1).

The association of lung function with blood pressure is difficult to interpret, mirroring the inconsistency that has been seen in large observational studies. Overall, our MR seems to suggests a causal effect of FEV1/FVC on blood pressure (Figure3B, Figures D-F in Additional File 1). ‘High blood pressure’ (systolic greater than 130mmHg or diastolic greater than 85mm Hg) has been associated with lower FEV1 and FVC in NHANES III (34), systolic blood pressure has been associated with lower FVC in the 2001 Korean National Health and Nutrition Survey (KNHNS) (35) and with COPD (defined by smoking status and airway obstruction) in the later fifth KNHNS V (36). One report suggests associations may be explained by the use of antihypertensive medication (37). Our analysis supports a genetic correlation between pulse pressure and FEV1, but none with diastolic or systolic pressure; while MR analysis shows a causal effect of FEV1/FVC on blood pressure (Figure 3) (37). More studies are needed to confirm and understand these associations.

Mendelian Randomisation analysis are a great tool to use large scale GWAS results to gain public health relevant insights, however one of the major limitations in MR is weak instrument bias, meaning the variant explains little variation of the exposure. To overcome this source of bias, we selected variants from large scale GWAS (Table 1) in combination with an a priori defined threshold of 5×10^-8^ (Table 2 in Additional File 2). Additionally, we performed a power analysis, which showed that this MR analysis was sufficiently powered (Figure G in Additional File 1).

Another major challenge in this MR study is pleiotropy, the potential for a SNP used as instrument for a risk factor to affect more than one phenotype. Due to this complexity in the present MR study, we relied on consistency of results of multiple MR methods with different assumptions as well as results from multivariable MR(18). We used very common IVW MR method to create a precise reference estimate which, however, is vulnerable to pleiotropy and extreme values. As second more robust method (Figure 3) we used weighted median based method, which is robust to outliers and works even if up to 50% of variants are invalid. Third method in use was mode-based estimation (MBE) which is another consensus-based method (Additional File 1 methods) with similar properties as weighted median method. MBE relies on so called Zero Modal Pleiotropy Assumption(38). That means that even if the group of valid instruments is only 40%, those will make the largest group of estimates within the distribution of ratio estimates and thus be driving the causal estimate. Our fourth estimate (Figure 3) originates from robust MR Egger method (Figure 3) and is very common in MR literature(14, 39, 40). It attempts to model pleiotropy under the InSIDE (Instrument strength independent of direct effect)(41), which assumes that pleiotropic effects need to be uncorrelated with each other. An assumption that may not be met by all traits in our analysis. Additionally, we performed sensitivity analysis applying outlier robust approaches and excluding potentially pleiotropic and/or invalid instruments from the analysis. These recently developed methods MR-PRESSO(21) and CONMIX(22) attempt to model pleiotropy in MR analysis and provide a measure of pleiotropy for each SNP. We excluded SNPs flagged by these methods re-analysed the data and observed generally lower P-values for the associations presented in the study (Table 3 in Additional File 2).

## Conclusions

In conclusion, we provide evidence for genetic correlations between BMI, CRP, T2D and coronary artery disease with FEV_1_, FVC and their ratio. These correlations reflect causal associations for the effects of BMI on all lung parameters and for T2D on FVC and FEV1/FVC. These associations are broadly independent from each other and of other metabolic traits with a small proportion of the effect of T2D and BMI on impaired lung function being mediated by serum CRP. There was evidence that FEV1/FVC ratio have a causal effect on blood pressure but not on the other tested cardio-metabolic traits. Our results strongly support efforts to reduce obesity and T2D as measures to improve lung function and lung health in the general population.

## Data Availability

The following summary statistics analysed in this study are publicly available: BMI, T2D, Blood lipid levels, Blood pressure, CAD.
The following analysed in this study are available upon request:
CRP: Summary statistics are available from the corresponding author on request.
UK Biobank data are available upon request (https://www.ukbiobank.ac.uk/register-apply/)
NFBC1966 data are available upon request (https://www.oulu.fi/nfbc/node/40677)

http://portals.broadinstitute.org/collaboration/giant/index.php/GIANT_consortium_data_files

http://diagram-consortium.org/downloads.html

http://lipidgenetics.org

https://www.ncbi.nlm.nih.gov/projects/gap/cgi-bin/study.cgi?study_id=phs000585.v1.p1

http://www.cardiogramplusc4d.org/data-downloads/

## List of abbreviations

BMI: body mass index
CAD: coronary artery disease
CRP: C-reactive protein
DBP: diastolic blood pressure
FEV1: forced expiratory volume in second one
FVC: forced vital capacity
FEV1/FVC: ratio of forced expiratory volume in second one and forced vital capacity
HDL-C: high density lipoprotein cholesterol
LDL-C: low density lipoprotein cholesterol
LD-score regression: linkage disequilibrium score regression
MR: Mendelian Randomisation
MMR: multivariable Mendelian Randomisation
PP: pulse pressure
SBP: systolic blood pressure
SNP: single nucleotide polymorphism
TC: total cholesterol
TG: triglycerides
T2D: type 2 diabetes

## Ethics approval

An informed consent for the use of the data including DNA was obtained from all participants of the Northern Finland Birth Cohort 1966.

## Availability of data and materials

The following summary statistics analysed in this study are publicly available:

BMI (http://portals.broadinstitute.org/collaboration/giant/index.php/GIANT_consortium_data_files);

T2D (http://diagram-consortium.org/downloads.html);

Blood lipid levels (http://lipidgenetics.org);

Blood pressure (https://www.ncbi.nlm.nih.gov/projects/gap/cgi-bin/study.cgi?study_id=phs000585.v1.p1);

CAD: (http://www.cardiogramplusc4d.org/data-downloads/).

The following analysed in this study are available upon request:

CRP: Summary statistics are available from the corresponding author on request.

UK Biobank data are available upon request (https://www.ukbiobank.ac.uk/register-apply/)

NFBC1966 data are available upon request (https://www.oulu.fi/nfbc/node/40677)

## Author contributions

MW prepared the first draft. MW, MRJ, and DLJ designed the study. MW, AA, SS, DvP and DMA analysed the data. All authors contributed to the interpretation of the data, participated in the drafting and critical revision of the manuscript, and granted final approval for submission.

## Acknowledgments

The authors are greatful to the late professor Paula Rantakallio (launch of NFBC1966), the participants in the 31- and 46-years-old study and the NFBC project center (www.oulu.fi/nfbc).

## Additional File 1 Methods

### Observational analyses in NFBC1966

The Northern Finland Birth Cohort 1966 (NFBC1966), described in detail previously (1, 2), targeted all pregnant women, residing in the two northernmost provinces of Finland with expected dates of delivery between 1 January and 31 December 1966. Over 96% of eligible women participated in the study, giving birth to 12,058 live born children. In 2012, at offspring age of 46 years, all cohort participants with known addresses and living in Northern Finland or Helsinki area were invited to a clinical examination, which included blood sampling. Clinical data and blood was collected from 5,861 participants. Lung function was assessed with a Vitalograph P spirometer (Vitalograph Ltd., Maids Moreton, UK). The present analysis is based on the best (highest) available lung function measure from participants who performed at least three acceptable blows, with the difference between two maximal readings of FEV1 or FVC less than 4% (3). Associations of lung function (FEV1, FVC, FEV1/FVC) with metabolic syndrome traits were investigated in linear regression models adjusted for sex (male/female), age (years), height (cm), smoking status (current, former, and never smokers) and pack-years (Appendix table 3).

Type 2 diabetes in NFBC1966 was defined as either or: prescription of metformin (Finnish register for reimbursed medication; ATC code A10B, available from year 1997 and 2016), diagnosed by a physician (Finnish outpatient register; ACD9 or 10) or screen-detected by OGTT at the age of 46y (NFBC1966 clinical follow-up in 2012). Coronary heart disease was defined based on participants answer to the following questions: “Do you now or have you had following the doctor diagnosed or treated the symptoms, diseases or injuries: Congenital heart disease”.

For Spirometry measurements, we used a MasterScreen Pneumo Spirometer (Vitalograph Ltd., Buckingham, UK), with a volumetric accuracy of ±2% or ±50 mL whichever was greater. The machines were calibrated every day the medical examination took place. The spirometric manoeuvre was performed three times in an upright sitting position while wearing a nose clip, but repeated if the coefficient of variation between two maximal readings was >4%.

### SNP-metabolic syndrome trait associations

We extracted the effect estimates for SNPs associated (P< 5×10-8) with BMI, T2D, CRP, blood lipids, blood pressure and CAD from publicly available lists. These summary statistics resulted from large meta-analyses of at least 80,000 individuals (table 1). The estimates for BMI-SNPs associations are from a GIANT consortium study (4), blood pressure associated SNPs are from a study by Wain et al. (5), blood lipid associated SNPs are from the Global Lipids Genetics consortium (6), CAD associated SNPs are from the Cardiogram consortium (7), T2D associated SNPs are from the DIAGRAM consortium (8), and CRP associated SNPs are from a study by Dehghan et al. (9). Individual summary statistics underwent study specific QC as well as genomic control procedure. Genomic coordinates were lifted over to GRCh37/hg19 if necessary.

### SNP-lung function associations

We used the UK Biobank release from July 2017. We restricted to HRC imputed SNPs due to possible wrong genomic position of non-HRC imputed SNPs. Genotyped SNPs were filtered for MAF > 5%, HWE P > 1×10-6, and missingness < 0.015, to estimate the kinship matrix. Genetic association results were retrieved for ~7.1 million SNPs remaining after filtering the HRC imputed data. The association tests were performed using linear mixed models (LMM) for all typed and imputed SNPs in dosage format using the BOLT-LMM (v2.3) software (10), which corrects for population structure and cryptic relatedness. We assumed an additive mode of inheritance.

Lung function measurements in the UK Biobank were generated with a Vitalograph Pneumotrac 6800 spirometer (Vitalograph Ltd., Maids Moreton, UK) according to recommendations. The present analysis is based on the best (highest) available lung function measure from those who performed at least two acceptable blows and where both FEV1 and FVC, although not from the same blow, were reproducible within 150 mL, as recommended by the American Thoracic Society/European Respiratory Society (ATS/ERS)(11).

### Cross trait LD score regression

All publicly available summary statistics (table 1) were corrected for genomic inflation. We used the LD score regression y-intercept as correction factor for genomic control procedure of the genome-wide association results obtained from the UK Biobank(12). We estimated pairwise genetic correlation and heritability using the recommended settings in LD score software (v1.0.0) (13).

### Mendelian Randomization

Mendelian Randomization relies on Mendel’s second law, that is, genotypes are assorted at random. This balances confounding and excludes reverse causation. A polymorphism associated with both the risk factor and with impaired lung function supports a causal role of the risk factor.(14). We performed a 2-sample Mendelian Randomization. If not indicated otherwise, MR analyses were performed using the CRAN package *Mendelianrandomization* (15).

#### IVW MR

We estimated the causal effect of the risk factor on lung function using widely used IVW MR. We combined inverse variance weighted ratios of genetic variants using a fixed effect meta-analysis model (16). We retrieved IVW Estimates and P values as well Cochran’s Q and a P value for heterogeneity as indication of Pleiotropy (Appendix table 6). Briefly, if all genes represent valid instruments, their MR estimates should vary only by chance. The presence of pleiotropy is then investigated by using the between-instrument heterogeneity Q test derived separately for each instrument(17). The P value tests the null hypothesis that all genetic variants are estimating the same causal parameter; rejection of the null is an indication that one or more variants may be pleiotropic. For IVW BMI effect size estimates, we performed linear regression analyses between the SNPs and BMI (N=453,868) adjusted for age, age^2^, centre, first 10 principal components (PCs) and genotyping batch (UK BiLEVE array and the UK Biobank Axiom array (2 data releases)).

### Weighted Median method

Similar to IVW method, however using the median of the ratio estimates as opposed to the mean in IVW. In the weighted version we add a weight corresponding to the precision of the to ratios. The method will provide unbiased estimates even if up to 50% of the variants are invalid instruments.

### Mode based estimation

The causal estimate is the maximum of a density function that is constructed from a normal density of each genetic variant. Similar to weighted median method this method is robust to outliers and has a higher break point as IVW and MR Egger method. MBE method is also less sensitive to violation of the InSIDE assumptions, because it relaxes the instrumental variable assumptions.

### MR Egger

MR Egger method attempts to model the distribution of invalid instruments. We regress the effect sizes of the variant-outcome associations against effect sizes of variant-exposure associations. An unconstrained interception term should reflect pleiotropy and removes the assumption that all instrumental variables are valid.

### Sensitivity analysis

We performed penalized versions for some of the methods mentioned above. Furthermore, we rerun the analysis with an altered set input SNPs based on outlier SNPs detected by either MR-PRESSO or the contamination mixture method. MR PRESSO was developed to detect horizontal pleiotropy. The variants effects on the outcome are regressed against the variant effects on the exposure. Again, regression line is the causal estimate. Then the same regression is performed without the tested variant and a residual sum of squares is calculated as the difference between regression lines. To obtain a P-value specific for pleiotropy of every variant a H0 distribution of RSS is simulated assuming no horizontal pleiotropy. Then we rerun IVW MR method without the variants flagged up by MR-PRESSO. Similar we run contamination mixture method to create a subset of SNPs to use in our sensitivity analysis. Contamination mixture method constructs a likelihood function based on the variant specific causal estimates if the instrument is valid the causal estimate of the tested variant will be around this estimate if not it will be around 0 with a high standard deviation. Based on the contribution of each variant to the likelihood the tested variant will be rated valid of invalid.

#### Multivariable MR

To account for the high genetic correlation between the different metabolic syndrome traits (Appendix Table 1), we used a recently developed extension of MR, called multivariable MR ^16^. We regressed the coefficients for the SNP-outcome association against all risk factors simultaneously and used the residuals of that regression as outcome to estimate the causal effect. We used the weighted regression-based approach to achieve this ^17^. We present raw P values. The Bonferroni threshold correcting for 18 tests (6 metabolic syndrome traits and 3 lung functions measurements) is 2.7×10-3.

#### Bidirectional MR

We repeated the MR in the opposite direction to determine possible causal effects of lung function on metabolic syndrome traits (Figure 1). For this we used a set of validated SNPs described by Wain et al (18) as instruments for lung function and performed a multivariable MR and IVW MR (Appendix methods, Appendix Table 1).

### Instrument strength and Power analysis

We calculated the cumulative variance explained applying the formula VarExp = beta^2 (1 - f) 2f, where beta is the coefficient for the SNP association to risk factor and f is the effect allele frequency. For this calculation as well as for Mendelian Randomization we included the most significant variant within a 1MB window out of a pool of variants with a P-value less than 5E-08. On a genome-wide level we calculated the genomic inflation factor lambda and extracted LD score intercept as indicator of the association between the risk or outcome and the genotype (Appendix table 3). Power calculations for our MR analysis were done specifically for binary and continuous outcomes (19, 20) The parameters for these calculations was the actual sample numbers of the outcome summary statistic, the cumulative variance explained as effect size of the risk factor and for the alpha level we used 2xE-03, the Bonferroni adjusted significance threshold used in this study. Power was calculated for varying true causal effect estimates ranging from 0.01 to 0.2. Power estimates at a true causal effect estimate of 0.1 are given Appendix table 4 and 5.

## Additional File 1 tables

**Additional File 1 table A:** Characteristics of summary statistics and Instrument strength; *validated loci used for invers direction (described by Wain et al. doi: 10.1038/ng.3787); **reported in study; ***European ancestry. Lambda GC values for lung function values are before genomic control correction. N SNPs is the number of SNPs reported as genome wide significant in published GWAS study.

| trait | source | lambda<br>GC | LDSC<br>intercept | N SNPs<<br>5xE-08 | cummulative<br>variance |
| --- | --- | --- | --- | --- | --- |
| FEV1 | UKBB & Wain et al. | 1.55 | 1.1 | 232 (32*) | 0.096** |
| FVC | UKBB & Wain et al. | 1.6 | 1.12 | 227 (21*) | 0.064** |
| FEV1/FVC | UKBB & Wain et al. | 1.55 | 1.1 | 332 (59*) | 0.143** |
| BMI | Locke et al. | 1.08 | 0.88 | 77*** | 0.027** |
| DBP | Wain et al. | 0.99 | 0.85 | 54 | 0.013 |
| SBP | Wain et al. | 1 | 0.87 | 55 | 0.013 |
| PP | Wain et al. | 1 | 0.87 | 31 | 0.005 |
| CAD | Nikpay et al. | 1.03 | 0.9 | 48 | 0.133** |
| CRP | Dehghan et al. | 1.09 | 1 | 18 | 0.05** |
| HDL | Willer et al. | 1.01 | 0.91 | 106 | 0.017 |
| LDL | Willer et al. | 1.01 | 0.98 | 82 | 0.023 |
| TG | Willer et al. | 1.01 | 0.98 | 66 | 0.013 |
| TC | Willer et al. | 1 | 0.96 | 102 | 0.023 |
| T2D | Scott et al. | 1.07 | 1.01 | 28 | 0.045 |

**Additional File 1 table B:** UK Biobank cohort characteristics (N = 270,381).

|  | % or mean (SD) |
| --- | --- |
| male | 42.8 |
| female | 57.1 |
| current smokers | 10.1 |
| Former smokers | 36.7 |
| Never smokers | 52.9 |
| Pack-years | 22.9 (18.0) |
| Age (years) | 56.6 (7.9) |
| Height (cm) | 1.68 (0.09) |
| FEV1 (L) | 2.81 (0.74) |
| FVC (L) | 3.71 (0.94) |
| FEV1/FVC | 0.76 (0.06) |

**Additional File 1 table C.** NFBC1966 cohort characteristics (N = 5567) for clinical examination at participants age 46.

|  | N (perc) or mean<br>(SD) |
| --- | --- |
| male | 2451 (44) |
| female | 3116 (55.9) |
| current smokers | 811 (15.4) |
| Former smokers | 2056 (38.9) |
| Never smokers | 2409 (45.6) |
| Pack-years | 10.4 (9.5) |
| Age (years) | 46.6 (0.6) |
| Height (cm) | 170.8 (9.1) |
| FEV1 (L) | 3.43 (0.71) |
| FVC (L) | 4.44 (0.96) |
| FEV1/FVC | 0.77 (0.06) |
| BMI | 26.8 (4.8) |
| CRP [mg/L] | 1.5 (2.8) |
| SBP | 128.8 (17.3) |
| DBP | 86 (11.2) |
| PP | 42.7 (10.4) |
| TC [mmol/L] | 5.3 (0.9) |
| TG [mmol/L] | 1.2 (0.8) |
| LDL [mmol/L] | 3.4 (0.6) |
| HDL [mmol/L] | 1.5 (0.3) |
| CAD cases | 136 (2.5) |
| CAD controls | 5128 (97.4) |
| T2D cases | 151 (3.1) |
| T2D controls | 4814 (96.9) |

**Additional File 1 table D:** Association between metabolic syndrome traits and lung function recorded in observational data retrieved from NFBC1966. Clinical assessment at participants age 46.

| Risk factor | Outcome | Estimate | SE | Pval | N |
| --- | --- | --- | --- | --- | --- |
| BMI | FVC | -0.019 | 0.002 | 9.73E-37 | 5293 |
| DBP | FVC | -0.004 | 0.001 | 1.18E-10 | 5287 |
| SBP | FVC | -0.002 | 0.000 | 4.93E-07 | 5287 |
| PP | FVC | -0.001 | 0.001 | 2.27E-01 | 5287 |
| CAD | FVC | -0.063 | 0.047 | 1.79E-01 | 5258 |
| CRP | FVC | -0.109 | 0.008 | 1.27E-45 | 4984 |
| HDL | FVC | 0.278 | 0.032 | 5.44E-18 | 5277 |
| LDL | FVC | -0.048 | 0.027 | 7.72E-02 | 5277 |
| TG | FVC | -0.174 | 0.016 | 2.99E-28 | 5277 |
| TC | FVC | -0.012 | 0.043 | 7.77E-01 | 5275 |
| T2D | FVC | -0.302 | 0.031 | 5.85E-22 | 5259 |
| BMI | FEV1 | -0.009 | 0.001 | 2.75E-13 | 5304 |
| DBP | FEV1 | -0.002 | 0.001 | 3.43E-03 | 5299 |
| SBP | FEV1 | -0.001 | 0.000 | 1.05E-02 | 5299 |
| PP | FEV1 | -0.001 | 0.001 | 3.15E-01 | 5299 |
| CAD | FEV1 | -0.034 | 0.038 | 3.67E-01 | 5270 |
| CRP | FEV1 | -0.069 | 0.006 | 4.16E-28 | 4993 |
| HDL | FEV1 | 0.140 | 0.026 | 1.16E-07 | 5286 |
| LDL | FEV1 | -0.007 | 0.022 | 7.62E-01 | 5286 |
| TG | FEV1 | -0.117 | 0.013 | 2.53E-19 | 5286 |
| TC | FEV1 | 0.004 | 0.035 | 9.17E-01 | 5284 |
| T2D | FEV1 | -0.184 | 0.026 | 1.02E-12 | 5271 |
| BMI | FEV1/FVC | 0.001 | 0.000 | 4.27E-13 | 5276 |
| DBP | FEV1/FVC | 0.0001 | 0.000 | 2.48E-06 | 5270 |
| SBP | FEV1/FVC | 0.0001 | 0.000 | 1.38E-03 | 5270 |
| PP | FEV1/FVC | 0.0001 | 0.000 | 9.31E-01 | 5270 |
| CAD | FEV1/FVC | 0.001 | 0.005 | 7.94E-01 | 5241 |
| CRP | FEV1/FVC | 0.004 | 0.001 | 1.46E-05 | 4968 |
| HDL | FEV1/FVC | -0.018 | 0.004 | 1.64E-06 | 5260 |
| LDL | FEV1/FVC | 0.008 | 0.003 | 1.19E-02 | 5260 |
| TG | FEV1/FVC | 0.004 | 0.002 | 2.27E-02 | 5260 |
| TC | FEV1/FVC | 0.003 | 0.005 | 5.26E-01 | 5258 |
| T2D | FEV1/FVC | 0.011 | 0.004 | 1.98E-03 | 5242 |

**Additional File 1 table E:**
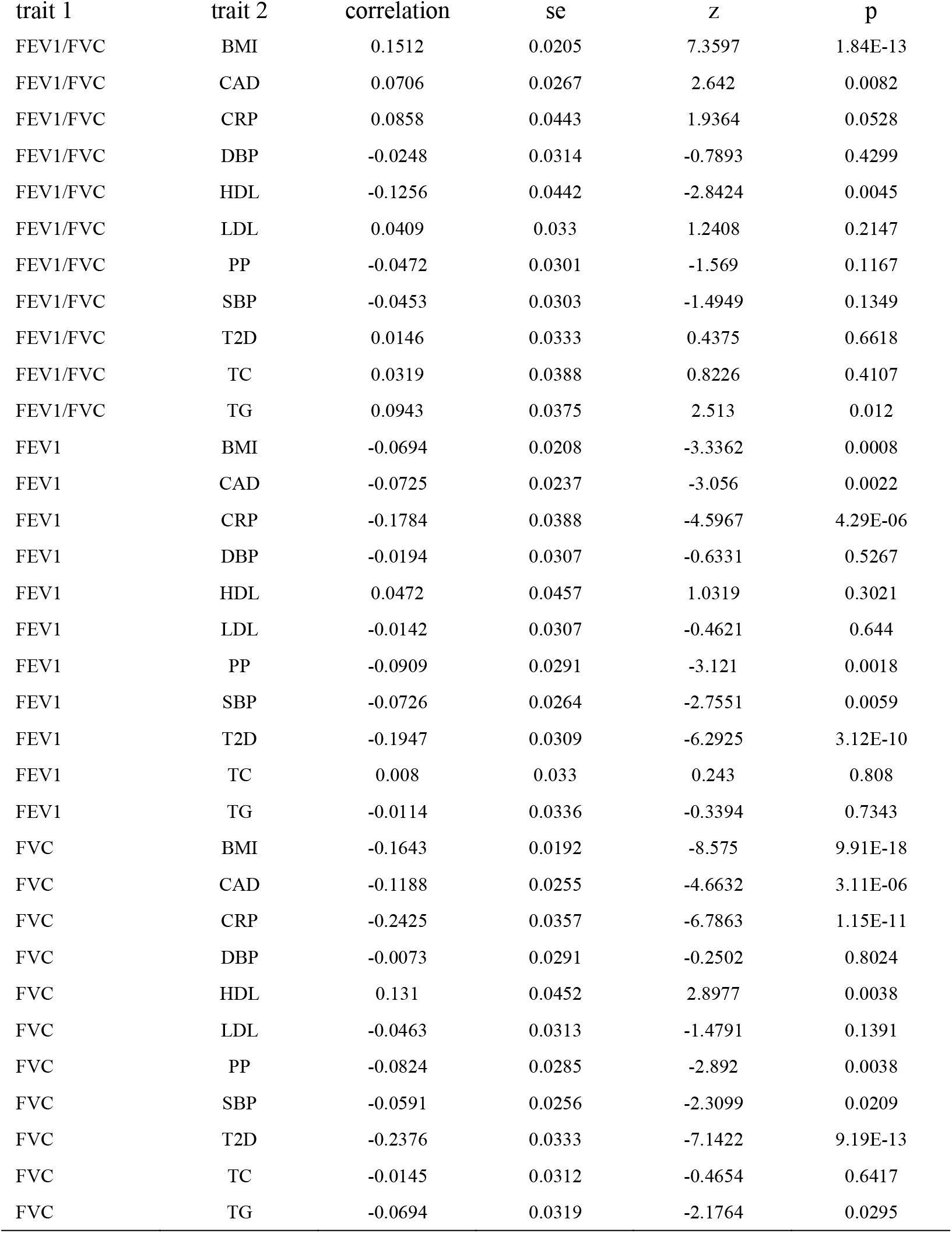
Result of cross trait LD score regression. Correlation values range from −1 to +1. Multiple testing threshold for this analysis is P < 0.0027.

| trait 1 | trait 2 | correlation | se | z | p |
| --- | --- | --- | --- | --- | --- |
| FEV1/FVC | BMI | 0.1512 | 0.0205 | 7.3597 | 1.84E-13 |
| FEV1/FVC | CAD | 0.0706 | 0.0267 | 2.642 | 0.0082 |
| FEV1/FVC | CRP | 0.0858 | 0.0443 | 1.9364 | 0.0528 |
| FEV1/FVC | DBP | -0.0248 | 0.0314 | -0.7893 | 0.4299 |
| FEV1/FVC | HDL | -0.1256 | 0.0442 | -2.8424 | 0.0045 |
| FEV1/FVC | LDL | 0.0409 | 0.033 | 1.2408 | 0.2147 |
| FEV1/FVC | PP | -0.0472 | 0.0301 | -1.569 | 0.1167 |
| FEV1/FVC | SBP | -0.0453 | 0.0303 | -1.4949 | 0.1349 |
| FEV1/FVC | T2D | 0.0146 | 0.0333 | 0.4375 | 0.6618 |
| FEV1/FVC | TC | 0.0319 | 0.0388 | 0.8226 | 0.4107 |
| FEV1/FVC | TG | 0.0943 | 0.0375 | 2.513 | 0.012 |
| FEV1 | BMI | -0.0694 | 0.0208 | -3.3362 | 0.0008 |
| FEV1 | CAD | -0.0725 | 0.0237 | -3.056 | 0.0022 |
| FEV1 | CRP | -0.1784 | 0.0388 | -4.5967 | 4.29E-06 |
| FEV1 | DBP | -0.0194 | 0.0307 | -0.6331 | 0.5267 |
| FEV1 | HDL | 0.0472 | 0.0457 | 1.0319 | 0.3021 |
| FEV1 | LDL | -0.0142 | 0.0307 | -0.4621 | 0.644 |
| FEV1 | PP | -0.0909 | 0.0291 | -3.121 | 0.0018 |
| FEV1 | SBP | -0.0726 | 0.0264 | -2.7551 | 0.0059 |
| FEV1 | T2D | -0.1947 | 0.0309 | -6.2925 | 3.12E-10 |
| FEV1 | TC | 0.008 | 0.033 | 0.243 | 0.808 |
| FEV1 | TG | -0.0114 | 0.0336 | -0.3394 | 0.7343 |
| FVC | BMI | -0.1643 | 0.0192 | -8.575 | 9.91E-18 |
| FVC | CAD | -0.1188 | 0.0255 | -4.6632 | 3.11E-06 |
| FVC | CRP | -0.2425 | 0.0357 | -6.7863 | 1.15E-11 |
| FVC | DBP | -0.0073 | 0.0291 | -0.2502 | 0.8024 |
| FVC | HDL | 0.131 | 0.0452 | 2.8977 | 0.0038 |
| FVC | LDL | -0.0463 | 0.0313 | -1.4791 | 0.1391 |
| FVC | PP | -0.0824 | 0.0285 | -2.892 | 0.0038 |
| FVC | SBP | -0.0591 | 0.0256 | -2.3099 | 0.0209 |
| FVC | T2D | -0.2376 | 0.0333 | -7.1422 | 9.19E-13 |
| FVC | TC | -0.0145 | 0.0312 | -0.4654 | 0.6417 |
| FVC | TG | -0.0694 | 0.0319 | -2.1764 | 0.0295 |

## Additional File 1 Figures

**Additional File 1 Figure A:**
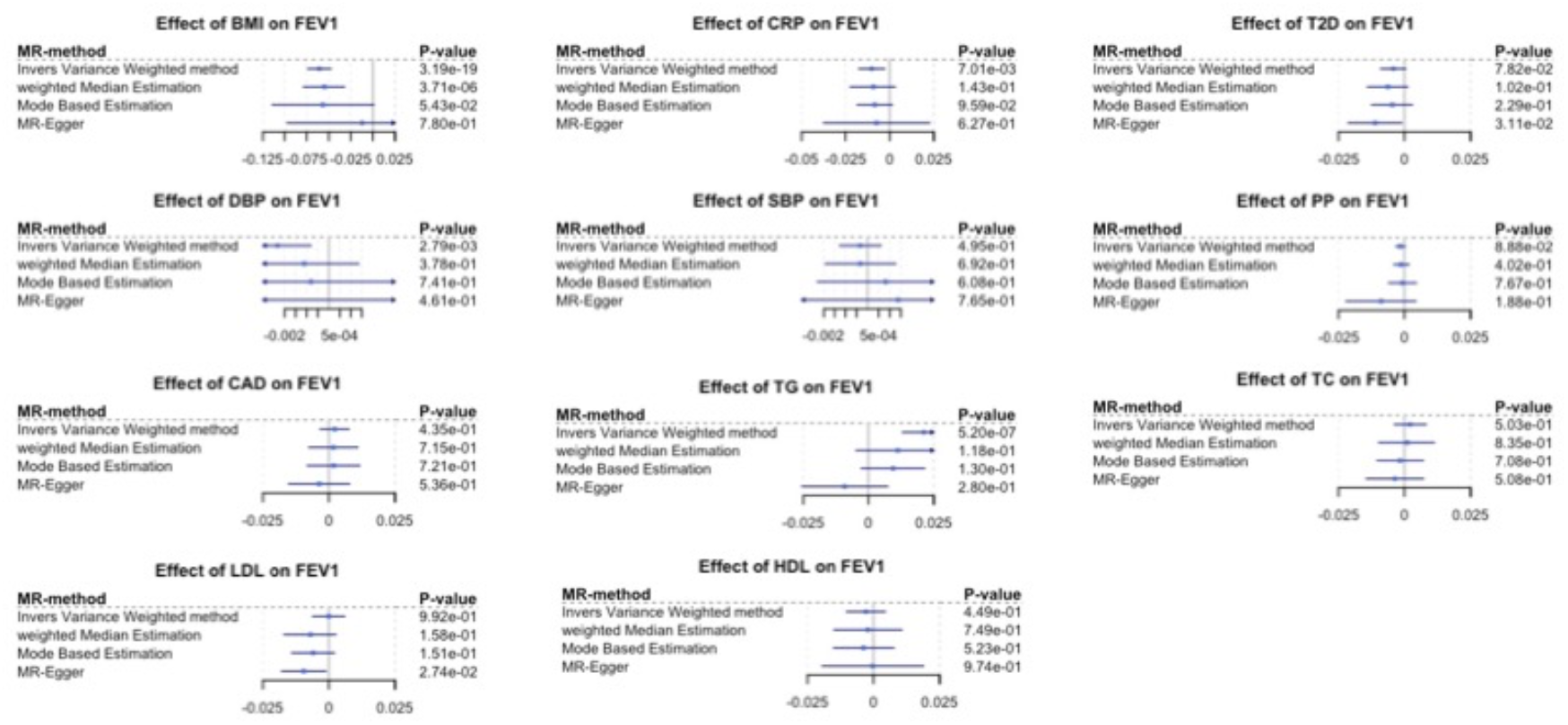
Forest plots of Metabolic Syndrome effects on FEV1. Blue square represents causal estimate. Blue line is 95% confidence interval. Every line represents one approach to estimate the potential causal effect.

**Additional File 1 Figure B:**
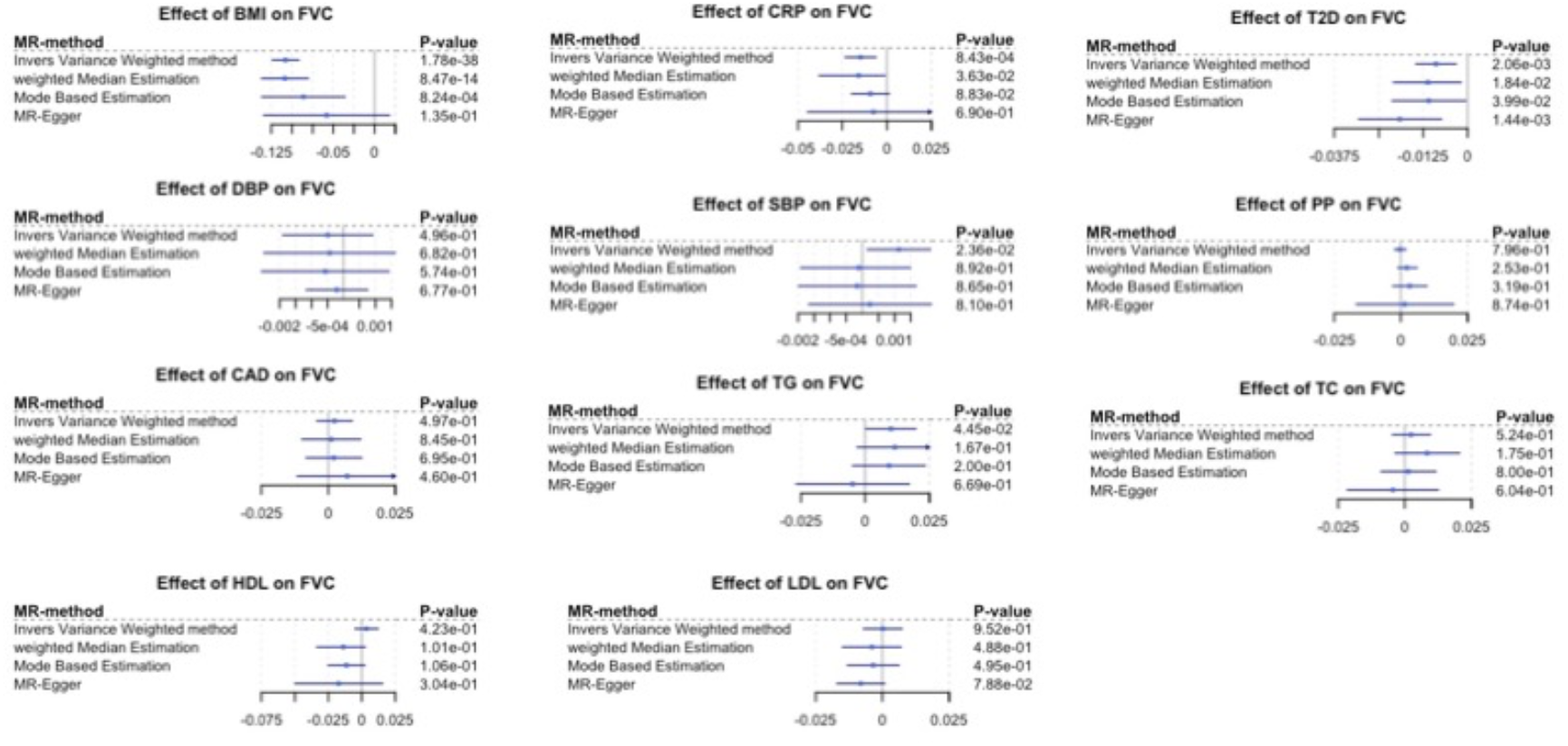
Forest plots of Metabolic Syndrome effects on FVC. Blue square represents causal estimate. Blue line is 95% confidence interval. Every line represents one approach to estimate the potential causal effect.

**Additional File 1 Figure C:**
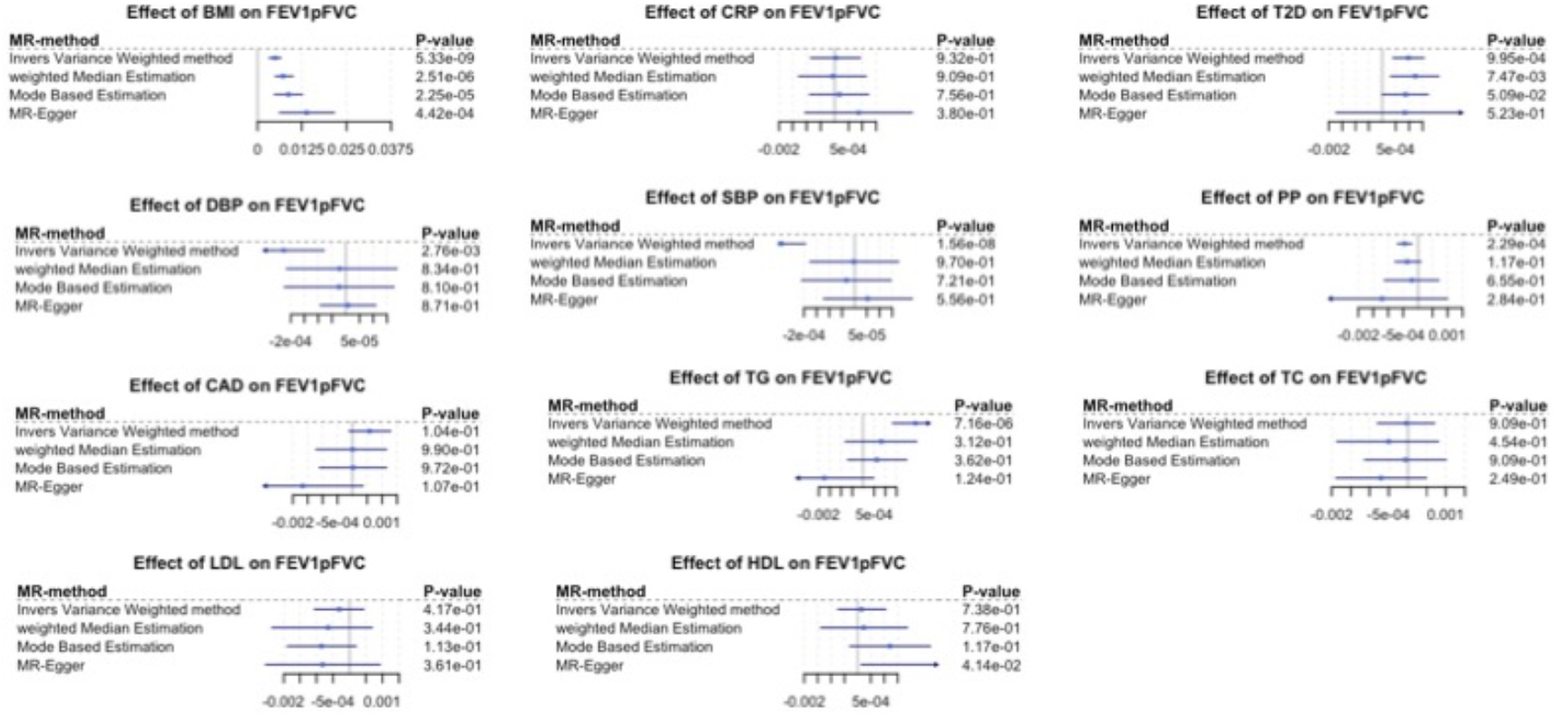
Forest plots of Metabolic Syndrome effects on FEV1pFVC. Blue square represents causal estimate. Blue line is 95% confidence interval. Every line represents one approach to estimate the potential causal effect.

**Additional File 1 Figure D:**
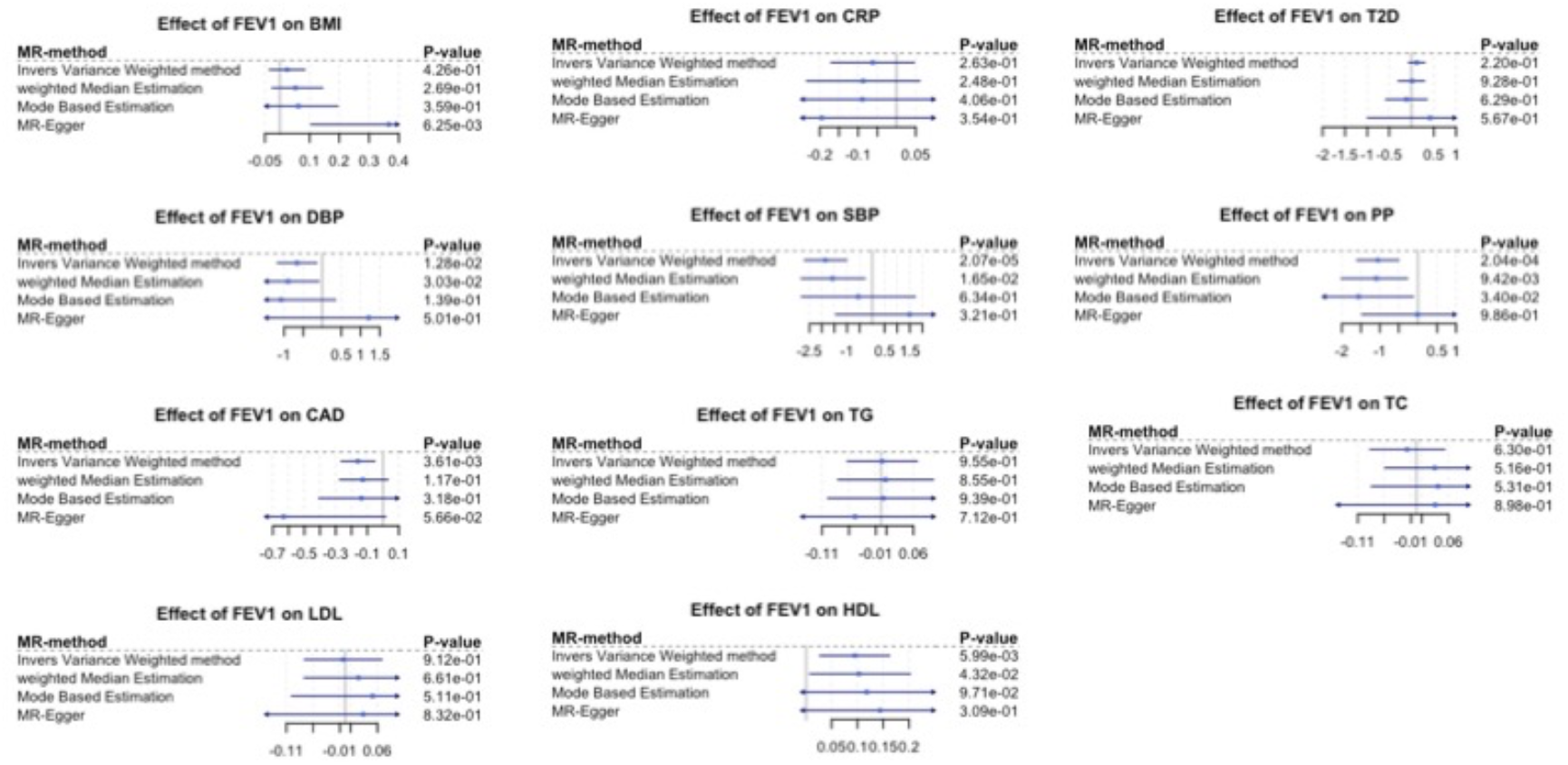
Forest plots of FEV1 effects on Metabolic Syndrome. Blue square represents causal estimate. Blue line is 95% confidence interval. Every line represents one approach to estimate the potential causal effect.

**Additional File 1 Figure E:**
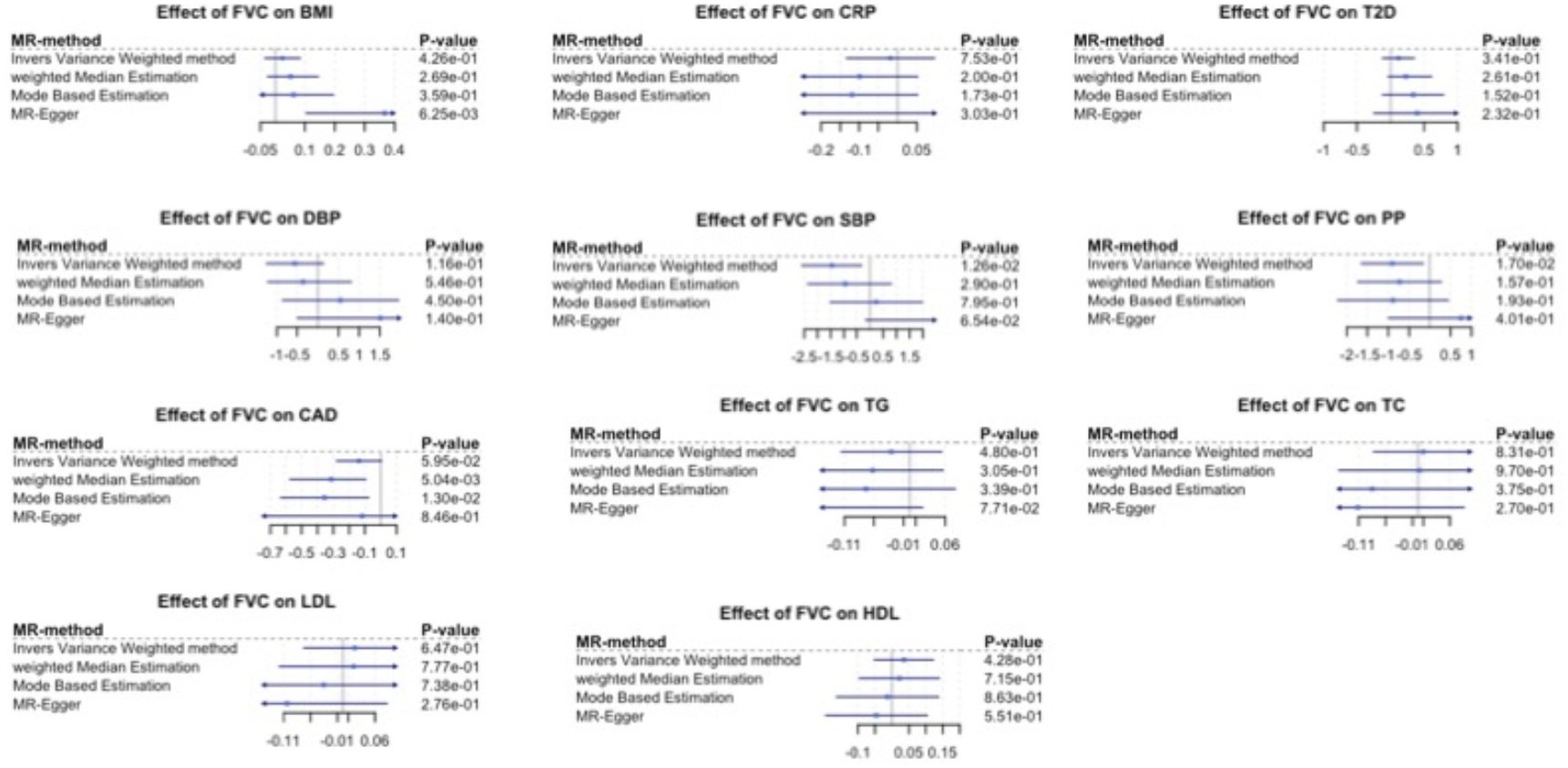
Forest plots of FVC effects on Metabolic Syndrome. Blue square represents causal estimate. Blue line is 95% confidence interval. Every line represents one approach to estimate the potential causal effect.

**Additional File 1 Figure F:**
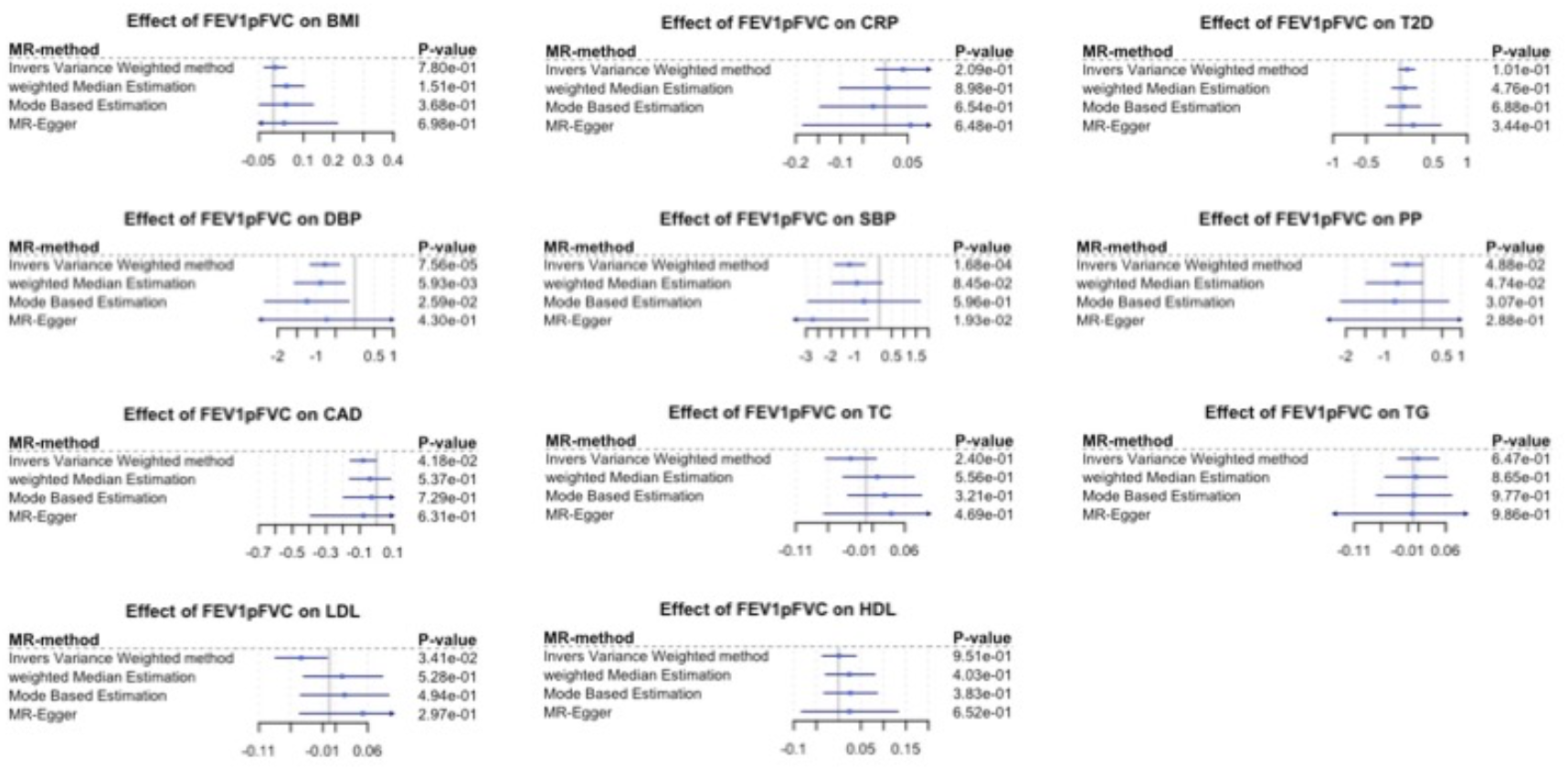
Forest plots of FEV1pFVC effects on Metabolic Syndrome. Blue square represents causal estimate. Blue line is 95% confidence interval. Every line represents one approach to estimate the potential causal effect.

**Additional File 1 Figure G:**
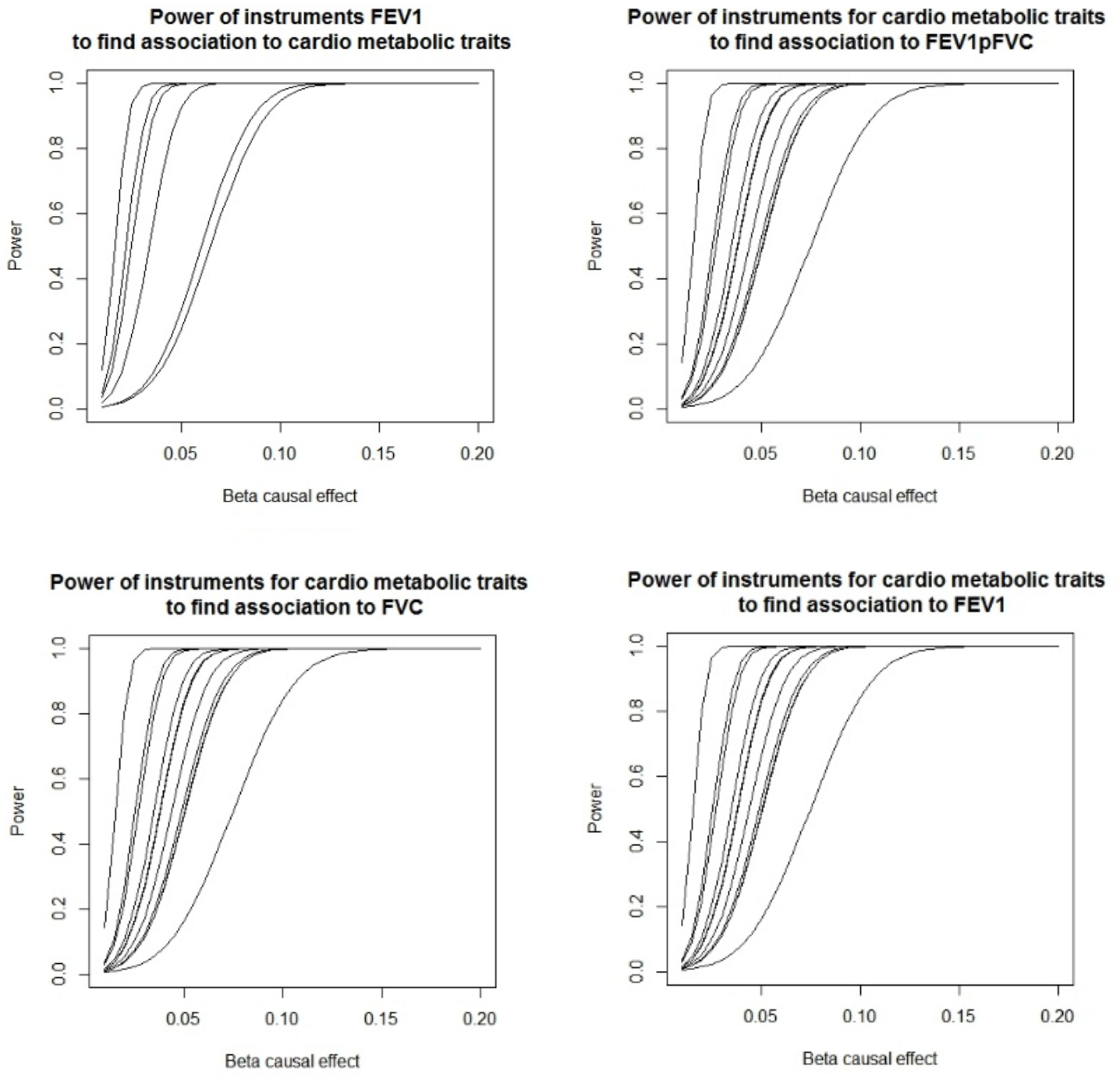
Power analysis. Where possible we used the variance explained reported by the studies, otherwise we calculated the variance explained of the PRS. We used an alpha level of 0.0028 and the actual sample numbers of the studies. For values at a causal effect estimate of 0.1.

